# Explainable Clinician-Supervised Artificial Intelligence as an Implementation Framework for Cardiovascular–Kidney–Metabolic Population Health: Synthetic Data Validation of the CHAPERONE-CKM Framework

**DOI:** 10.64898/2026.08.17.26360643

**Authors:** Aditya Vijay, Navin Govind, Arjun Moorthy, Patrick Dunn, Zaki Lababidi, Sun Jones, Marcus Stahlberg, Samer Ibrahim, Keyvan Koochek, Kevin Shah, Randal Schulhauser, Edgar V. Lerma, Lakshmi Nair, Jordi Livi, Dinesh Kalra, Devendra Wadwekar, Wes Gullett, Krishnaswami Vijayaraghavan

## Abstract

**Background:** Cardiovascular–kidney–metabolic (CKM) syndrome is an increasingly prevalent multisystem condition associated with morbidity, fragmented care, recurrent hospitalization, and rising healthcare costs. While cardiovascular risk models estimate future disease risk, fewer frameworks support multidisciplinary CKM care, clinician decision-making, and population health management. Synthetic data environments can assess implementation readiness while preserving privacy.

**Methods:** We validated the explainable, clinician-supervised CHAPERONE-CKM framework using a reproducible synthetic cohort of 10,090 simulated patients with 128 demographic, laboratory, imaging, treatment, and healthcare utilization variables across the CKM continuum. Synthetic data generation was separated from framework evaluation through probabilistic modeling and independent validation to reduce deterministic relationships. The framework generated CKM stage assignments, implementation priorities, clinician-readable rationales, multidisciplinary referral pathways, and guideline-directed therapy prompts. Evaluation focused on implementation readiness, consistency, calibration, subgroup stability, fairness, workflow simulation, and explainability.

**Results:** The synthetic population represented CKM-related conditions including diabetes (52%), hypertension (65%), chronic kidney disease (20%), heart failure (32%), and prior CKM hospitalization (27%). The framework showed stable internal behavior across demographic and clinical subgroups, favorable calibration, and biologically plausible prioritization of advanced CKM disease. Workflow simulations suggested earlier identification of patients suitable for multidisciplinary review, therapy optimization, and coordinated care compared with reactive workflows. Traditional performance metrics supported framework behavior but were treated as secondary evidence rather than proof of clinical effectiveness.

**Conclusions:** In a synthetic validation environment, the CHAPERONE-CKM framework demonstrated implementation readiness, transparent decision pathways, and compatibility with multidisciplinary CKM population health management. These findings are an early translational milestone, not clinical validation, and support external validation, prospective implementation studies, and Learning Health System integration to assess effects on care delivery, equity, and value-based outcomes.

## Introduction

Cardiovascular–kidney–metabolic (CKM) syndrome has emerged as a unifying construct recognizing the interconnected progression of obesity, diabetes mellitus, chronic kidney disease (CKD), atherosclerotic cardiovascular disease (ASCVD), and heart failure. Rather than representing isolated disorders, these conditions evolve through shared metabolic, inflammatory, neurohormonal, and vascular pathways that collectively accelerate morbidity, mortality, and healthcare utilization. Recognition of this continuum has shifted emphasis from organ-specific management toward integrated prevention and multidisciplinary care across the CKM spectrum.^1–3^. This implementation-focused paradigm has been further reinforced by the 2026 AHA/ACC/ADA/ASN Guideline for Cardiovascular–Kidney–Metabolic Syndrome, which emphasizes interdisciplinary care teams, coordinated implementation of evidence-based therapies, comprehensive medication management, and longitudinal care pathways as foundational elements of CKM management across the disease continuum.^49^ These recommendations underscore the need for implementation frameworks that facilitate team-based care while preserving clinician oversight.

Despite major advances in evidence-based therapies, including sodium-glucose cotransporter-2 inhibitors, glucagon-like peptide-1 receptor agonists, nonsteroidal mineralocorticoid receptor antagonists, and contemporary heart failure therapies, implementation remains inconsistent. Patients with CKM syndrome continue to experience delayed diagnosis, fragmented specialty care, therapeutic inertia, and recurrent hospitalization despite therapies that substantially improve cardiovascular and renal outcomes.^3–7^ These implementation gaps disproportionately affect socioeconomically disadvantaged populations and contribute substantially to healthcare expenditures.

Recent cardiovascular risk equations, including the American Heart Association PREVENT equations, have improved long-term cardiovascular risk estimation by incorporating broader cardiometabolic determinants than earlier models.^4^ However, these tools were designed primarily for risk prediction and not to facilitate multidisciplinary CKM care, prioritize patients for coordinated intervention, or support implementation of guideline-directed medical therapy in individuals with complex multimorbidity.^5–10^

Artificial intelligence (AI) has demonstrated considerable promise in cardiovascular medicine through imaging interpretation, risk prediction, electronic health record analytics, and clinical decision support. However, widespread implementation remains limited by concerns regarding transparency, interpretability, governance, algorithmic bias, workflow integration, and clinician trust. Increasingly, implementation science recognizes that analytical performance alone is insufficient; successful deployment requires explainability, human oversight, seamless workflow integration, and continuous evaluation within Learning Health Systems.^11–12^

Explainable AI addresses these barriers by providing clinician-readable reasoning that augments rather than replaces professional judgment. In CKM care, where management requires coordination among primary care physicians, cardiologists, nephrologists, endocrinologists, pharmacists, nurses, and care coordinators, transparent decision support can facilitate patient prioritization, multidisciplinary referral, and optimization of evidence-based therapies.

Synthetic data have emerged as an important methodology for early evaluation of healthcare AI systems. By generating realistic but non-identifiable patient populations, synthetic datasets enable assessment of framework behavior, implementation workflows, calibration, and explainability while preserving patient privacy.^20–23^ Although synthetic validation cannot establish clinical effectiveness or replace external validation, it provides an efficient translational step for iterative refinement before prospective evaluation.

The **CHAPERONE-CKM** framework was developed as an explainable, clinician-supervised implementation platform designed to support multidisciplinary CKM population health rather than function solely as a predictive model. The framework integrates multidomain cardiovascular, renal, metabolic, laboratory, therapeutic, and healthcare utilization data to generate transparent implementation prioritization with clinician-readable rationale while maintaining human oversight.

Accordingly, the objective of the present study was not to establish clinical efficacy but to evaluate implementation readiness within a synthetic CKM environment. We assessed the framework’s internal consistency, calibration, explainability, subgroup stability, fairness across demographic groups, and compatibility with multidisciplinary workflows. We hypothesized that synthetic data validation would demonstrate a reproducible implementation framework suitable for progression to external validation and prospective evaluation within Learning Health Systems.

## Methods

### Study Design

CHAPERONE-CKM is a prospective multiphase translational investigation evaluating an explainable artificial intelligence–enabled ecosystem for cardiovascular–kidney– metabolic (CKM) management. The program comprises sequential phases including early response evaluation, synthetic framework validation, device validation, feasibility testing, and pragmatic registry evaluation. Phase Ia, presented at the American Heart Association Scientific Sessions 2025, assessed the correctness, trustworthiness, usability, empathy, comprehensiveness, and clinical relevance of AI-generated responses. Phase Ib, the focus of this study, evaluated the CHAPERONE-CKM implementation framework using a reproducible synthetic cohort. Phase IIa is evaluating CKMiq, a noninvasive impedance-based hemodynamic assessment device against simultaneous echocardiography, followed by a feasibility study (Phase IIb) involving 30 participants recently hospitalized for acute CKM events. Phase III will evaluate the integrated platform in a pragmatic registry of approximately 1,200 participants.

Phase Ib employed a synthetic data validation design to assess implementation readiness before prospective deployment. The objective was not to establish diagnostic accuracy or clinical effectiveness but to determine whether an explainable clinician-supervised framework demonstrated coherent implementation behavior, transparent reasoning, subgroup stability, and compatibility with multidisciplinary CKM workflows. Findings are interpreted as early translational validation rather than evidence of real-world clinical performance.

### CHAPERONE-CKM Framework

CHAPERONE-CKM was developed as an explainable clinician-supervised implementation framework intended to augment—not replace—clinical judgment. The framework integrates demographic characteristics, cardiovascular risk factors, kidney function, metabolic variables, laboratory measurements, imaging findings, therapeutic information, and healthcare utilization data to generate CKM stage assignment, implementation priority, composite hazard estimation, clinician-readable explanations, multidisciplinary referral recommendations, and opportunities to optimize guideline-directed medical therapy. Framework design was informed by implementation science, Learning Health Systems, and responsible AI principles while preserving clinician authority for all diagnostic and therapeutic decisions.

### Synthetic Data Generation

A reproducible synthetic cohort of **10,090 virtual patients** representing the CKM continuum was generated using probabilistic methods with biologically plausible conditional dependencies derived from published epidemiologic evidence. The dataset comprised **128 demographic, laboratory, imaging, therapeutic, and healthcare utilization variables**, of which approximately **60 multidomain clinical variables** were integrated by CHAPERONE-CKM during framework evaluation.

Synthetic cohort generation was intentionally separated from downstream CKM adjudication to minimize deterministic relationships and label leakage. Automated physiological validation assessed concordance among estimated glomerular filtration rate, CKD stage, diabetes status, albuminuria, natriuretic peptide concentrations, and heart failure phenotype. Records violating predefined biological constraints were regenerated before analysis. Complete computational specifications are provided in **Supplementary Methods S1 and S13**.

### Framework Outputs and Workflow Simulation^13-20,34-38^

For each synthetic individual, CHAPERONE-CKM generated CKM stage, implementation priority, composite hazard estimate, clinician-readable explanations, multidisciplinary referral recommendations, and opportunities for optimization of guideline-directed medical therapy. These outputs were designed to support population health management and multidisciplinary clinical decision-making rather than autonomous diagnosis or treatment selection.

Implementation feasibility was evaluated within a simulated electronic health record– enabled workflow involving primary care, cardiology, nephrology, endocrinology, pharmacy, nursing, and care coordination. Framework recommendations supported referral prioritization, identification of high-risk individuals, and optimization of evidence-based CKM care. The implementation strategy was aligned conceptually with the RE-AIM and Consolidated Framework for Implementation Research (CFIR); detailed workflow specifications are provided in **Supplementary Methods S16**.

### Outcome Measures and Analyses

Primary outcomes evaluated implementation readiness, including internal consistency, transparency of clinician-readable explanations, workflow compatibility, subgroup stability, fairness, and suitability for Learning Health System integration.^121-30^ Traditional statistical measures—including calibration, discrimination, decision-curve analysis, and net reclassification improvement—were considered secondary implementation metrics supporting interpretation of framework behavior rather than estimates of clinical performance.

Prespecified subgroup analyses examined age, sex, race and ethnicity, diabetes status, CKD stage, and heart failure phenotype. Because all records were synthetic, subgroup analyses assessed implementation consistency rather than healthcare disparities.

### Statistical Analysis

Continuous variables are presented as means with standard deviations or medians with interquartile ranges, and categorical variables as frequencies and percentages. Calibration was evaluated using calibration plots, calibration slope, calibration intercept, the Hosmer–Lemeshow statistic, and the Brier score. Discrimination was assessed using the area under the receiver operating characteristic curve with 95% confidence intervals. Decision-curve analysis examined potential implementation utility across clinically relevant thresholds, while net reclassification improvement was calculated relative to a simplified PREVENT-aligned comparator. Internal stability was evaluated using **1,000 bootstrap resamples** to estimate optimism-adjusted implementation metrics.

### Reporting Standards

The study followed contemporary reporting recommendations for AI-enabled clinical research, including **TRIPOD+AI, CONSORT-AI, SPIRIT-AI, DECIDE-AI, MINIMAR,** and **StaRI**.^34-38^ Framework governance emphasized transparency, clinician oversight, reproducibility, auditability, and responsible implementation. Detailed reporting checklists, governance documentation, interoperability specifications, and mathematical methods are provided in the **Supplementary Appendix**.

## RESULTS

### Characteristics of the Synthetic CKM Population

The synthetic validation cohort comprised **10,090 individuals** representing the full spectrum of cardiovascular–kidney–metabolic disease. Demographic characteristics and disease prevalence were designed to reflect the heterogeneity encountered in contemporary multidisciplinary practice. More than one-half of the cohort had diabetes mellitus (52%), nearly two-thirds had hypertension (65%), approximately one-third had established heart failure (32%), one-fifth had advanced chronic kidney disease (20%), and over one-quarter had experienced a previous CKM-related hospitalization (27%) (Table 1).

**Table 1.**
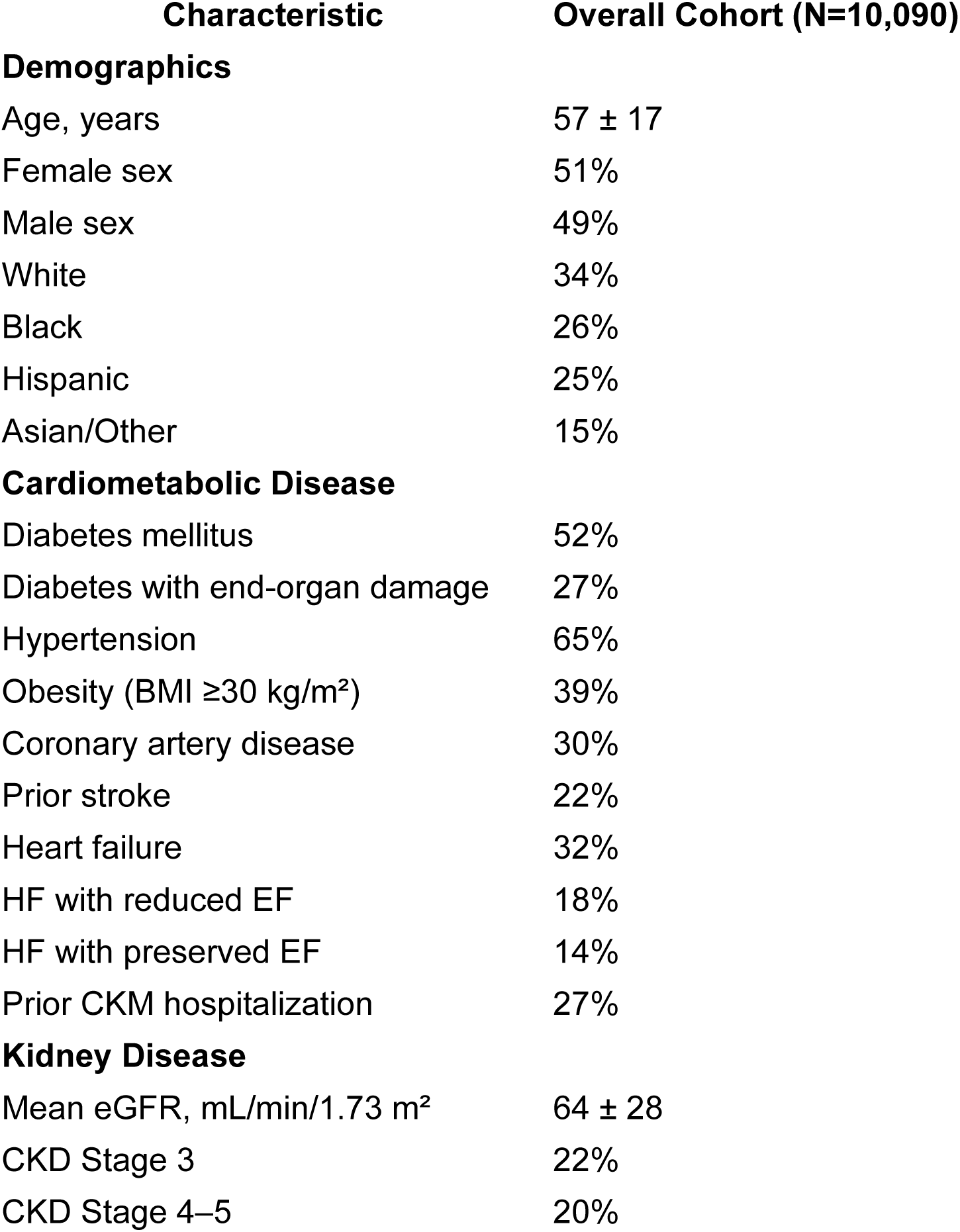
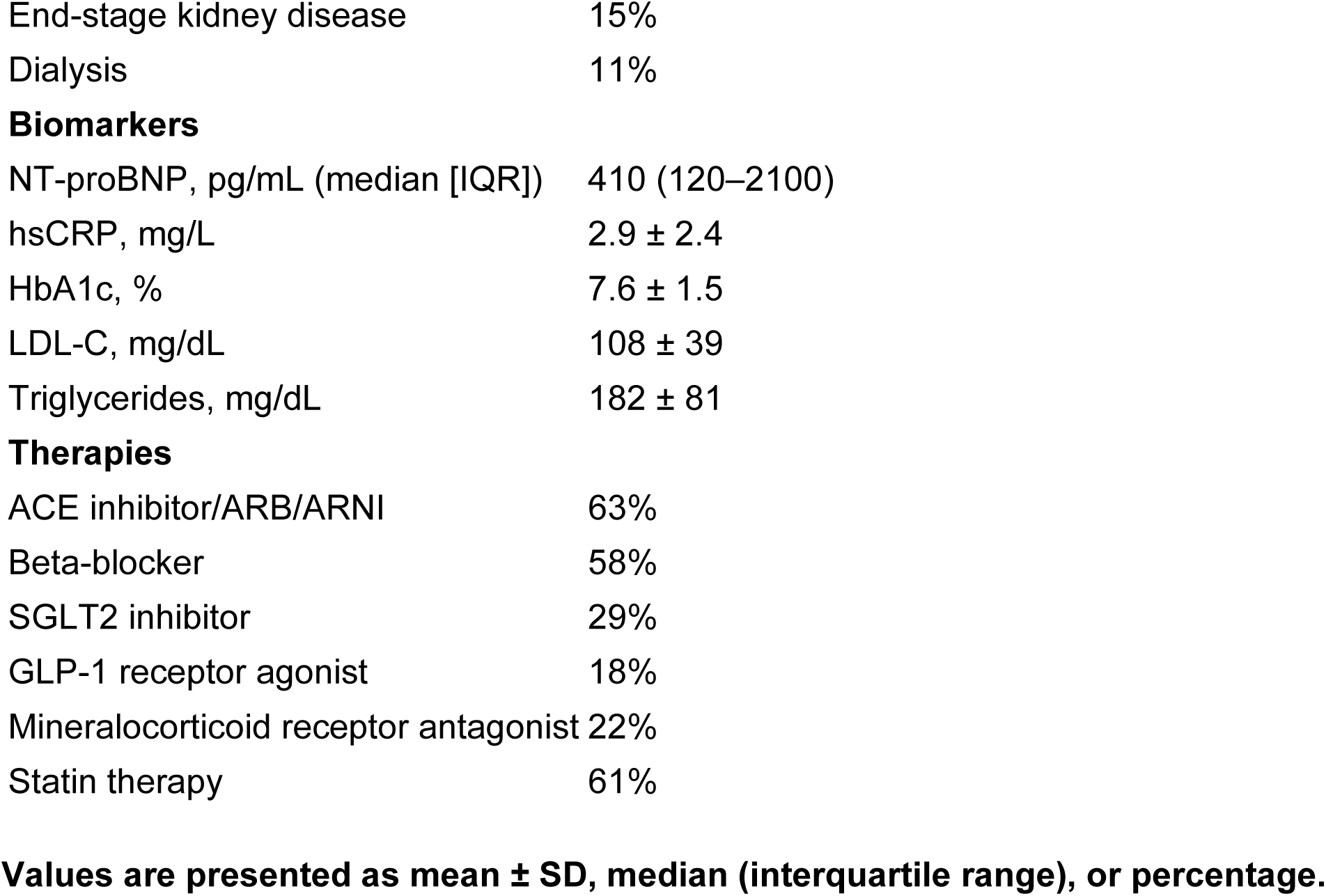
Baseline Characteristics of the Synthetic Cardiovascular–Kidney– Metabolic (CKM) Validation Cohort.

| Characteristic | Overall Cohort (N=10,090) |
| --- | --- |
| **Demographics** |  |
| Age, years | 57 ± 17 |
| Female sex | 51% |
| Male sex | 49% |
| White | 34% |
| Black | 26% |
| Hispanic | 25% |
| Asian/Other | 15% |
| **Cardiometabolic Disease** |  |
| Diabetes mellitus | 52% |
| Diabetes with end-organ damage | 27% |
| Hypertension | 65% |
| Obesity (BMI ≥30 kg/m<sup>2</sup>) | 39% |
| Coronary artery disease | 30% |
| Prior stroke | 22% |
| Heart failure | 32% |
| HF with reduced EF | 18% |
| HF with preserved EF | 14% |
| Prior CKM hospitalization | 27% |
| **Kidney Disease** |  |
| Mean eGFR, mL/min/1.73 m<sup>2</sup> | 64 ± 28 |
| CKD Stage 3 | 22% |
| CKD Stage 4–5 | 20% |
| End-stage kidney disease | 15% |
| Dialysis | 11% |
| **Biomarkers** |  |
| NT-proBNP, pg/mL (median [IQR]) | 410 (120–2100) |
| hsCRP, mg/L | 2.9 ± 2.4 |
| HbA1c, % | 7.6 ± 1.5 |
| LDL-C, mg/dL | 108 ± 39 |
| Triglycerides, mg/dL | 182 ± 81 |
| **Therapies** |  |
| ACE inhibitor/ARB/ARNI | 63% |
| Beta-blocker | 58% |
| SGLT2 inhibitor | 29% |
| GLP-1 receptor agonist | 18% |
| Mineralocorticoid receptor antagonist | 22% |
| Statin therapy | 61% |

**Values are presented as mean ± SD, median (interquartile range), or percentage.**

The cohort included broad representation across age, sex, race and ethnicity, cardiovascular disease severity, kidney dysfunction, metabolic burden, and healthcare utilization, providing a diverse environment for evaluating framework behavior across clinically relevant CKM profiles.

### Framework Behavior Within a Simulated CKM Care Environment

CHAPERONE-CKM demonstrated consistent implementation behavior across the synthetic CKM continuum (Table 2). Rather than functioning as an isolated prediction model, the framework integrated demographic characteristics, laboratory measurements, imaging findings, therapeutic information, and healthcare utilization into clinician-readable implementation prioritization supporting multidisciplinary care.

**Table 2.** Implementation Readiness Domains Evaluated During Synthetic Data Validation.

| Domain | Framework Objective | Primary Evaluation |
| --- | --- | --- |
| Explainability | Transparent clinician-readable recommendations | Explanation generated for every recommendation |
| Clinical Governance | Human oversight | Clinician-supervised framework |
| Population Health | CKM prioritization | High-risk patient identification |
| Workflow Integration | Multidisciplinary care coordination | Simulated referral pathway |
| Learning Health System | Continuous quality improvement | EHR implementation roadmap |
| Responsible AI | Safety and transparency | Auditability and deterministic inference |
| Framework Validation | Internal consistency | Calibration and discrimination |

Patients assigned higher implementation priority generally demonstrated advanced cardiorenal dysfunction, progressive metabolic disease, previous hospitalization, or combinations of these conditions. Every prioritization was accompanied by transparent explanations identifying the principal clinical contributors, preserving clinician interpretability and facilitating multidisciplinary review.

Within the simulated workflow, the framework consistently directed patients toward appropriate care pathways, including primary care, preventive cardiology, nephrology, endocrinology, heart failure management, or multidisciplinary CKM review according to the overall disease profile rather than isolated disease-specific variables (Figure 3).

### Implementation Workflow Simulation

Simulation within a Learning Health System demonstrated the potential for proactive CKM management through continuous electronic health record surveillance and dynamic implementation prioritization (Table 3). The workflow incorporated clinician review, care coordination, specialty referral, and reassessment following therapeutic optimization.

**Table 3.** Implementation-Oriented Clinical Workflow Simulation.

| Workflow Component | Conventional Care | CHAPERONE-CKM Framework |
| --- | --- | --- |
| Population surveillance | Periodic review | Continuous surveillance |
| Patient prioritization | Manual clinician review | Automated clinician-supported prioritization |
| CKM risk integration | Organ-specific | Multidomain CKM |
| Specialist referral | Reactive | Earlier multidisciplinary referral |
| GDMT assessment | Variable | Structured review prompts |
| Care coordination | Fragmented | Coordinated multidisciplinary workflow |
| Longitudinal monitoring | Episodic | Continuous population management |
| Clinician oversight | Standard | Preserved human oversight |

Across simulated scenarios, the framework consistently identified opportunities for optimization of guideline-directed medical therapy, multidisciplinary referral, reassessment of cardiovascular and kidney risk, and longitudinal monitoring within CKM population registries (Figure 4).

### Explainability and Clinical Transparency

Every implementation recommendation included a structured explanation identifying the principal digital biomarkers contributing to prioritization. Advanced chronic kidney disease, elevated natriuretic peptide concentrations, diabetes with end-organ injury, established ASCVD, previous CKM hospitalization, and systemic inflammatory markers consistently emerged as dominant contributors to higher implementation priority. Importantly, the framework generated implementation support rather than autonomous diagnostic or therapeutic recommendations, thereby preserving clinician oversight and shared decision-making.

### Supporting Framework Validation

Calibration and discrimination analyses were performed as secondary measures of framework behavior (Tables 4 and 5). Agreement between framework-generated CKM stage assignments and independently generated synthetic reference classifications supported internal consistency of implementation logic. Calibration analyses demonstrated favorable agreement between predicted and simulated outcomes, while discrimination analyses effectively separated lower- and higher-priority synthetic CKM profiles.

Decision-curve analysis suggested potential implementation utility across clinically relevant thresholds. Compared with a simplified PREVENT-aligned comparator, incorporation of multidomain cardiovascular, kidney, metabolic, and healthcare utilization variables improved identification of individuals with advanced synthetic CKM burden. Because all analyses were performed within a synthetic validation environment, these findings should be interpreted as evidence of framework behavior rather than clinical performance.

### Subgroup Consistency and Fairness Assessment

Framework behavior remained stable across prespecified subgroups, including age, sex, race and ethnicity, diabetes status, chronic kidney disease stage, and heart failure phenotype (Table 6). Calibration characteristics and implementation prioritization showed no evidence of systematic instability.

Fairness analyses likewise demonstrated consistent implementation behavior across demographic groups within the synthetic environment. Although these observations cannot establish equity in clinical practice, they support progression to prospective external validation in real-world populations.

### Learning Health System Readiness

The simulated implementation environment demonstrated compatibility with Learning Health System principles through integration with longitudinal electronic health record surveillance, multidisciplinary referral pathways, population health dashboards, and continuous quality improvement processes (Figure 5). The explainable architecture supported clinician review while maintaining transparency, auditability, and governance required for future health system deployment.

### Key Findings

Synthetic data validation demonstrated that an explainable clinician-supervised implementation framework can integrate multidomain CKM information into transparent patient prioritization while supporting multidisciplinary workflows, Learning Health Systems, and responsible artificial intelligence principles. Traditional statistical measures confirmed internal framework behavior but served as secondary implementation metrics. Collectively, these findings establish a translational foundation for external validation and prospective implementation studies evaluating the impact of clinician-supervised explainable AI on CKM care delivery and value-based outcomes.

## DISCUSSION

### Principal Findings

This study evaluated CHAPERONE-CKM within a synthetic validation environment as an early translational step toward cardiovascular–kidney–metabolic (CKM) population health implementation. Rather than developing another predictive algorithm, we investigated whether explainable, clinician-supervised artificial intelligence could function as an implementation framework supporting multidisciplinary care, transparent clinical reasoning, and Learning Health System integration. The framework demonstrated biologically coherent implementation prioritization, clinician-readable explanations, stable behavior across clinically relevant subgroups, and compatibility with simulated multidisciplinary workflows. These findings support implementation readiness within a synthetic environment but should not be interpreted as evidence of clinical effectiveness.

### Artificial Intelligence as an Implementation Framework

Most AI applications in cardiovascular medicine emphasize prediction, diagnosis, or image interpretation. Although these approaches have demonstrated strong analytical performance, clinical adoption has often been limited by inadequate explainability, workflow integration, and clinician trust. CHAPERONE-CKM was intentionally designed as an implementation framework that integrates cardiovascular, kidney, metabolic, laboratory, imaging, therapeutic, and healthcare utilization data into transparent recommendations supporting multidisciplinary care. This implementation focus is particularly relevant for CKM syndrome, where effective management requires coordinated care across specialties rather than isolated disease-specific decisions.

### Addressing Fragmentation in CKM Care

The American Heart Association CKM framework recognizes the biological interconnectedness of obesity, diabetes, CKD, ASCVD, and heart failure, yet healthcare delivery remains largely organ-specific. Fragmented care contributes to therapeutic inertia, duplicated testing, and delayed implementation of guideline-directed therapies. CHAPERONE-CKM addresses these gaps by synthesizing multi-domain digital biomarkers into clinician-readable implementation priorities that facilitate communication among primary care physicians, cardiologists, nephrologists, endocrinologists, pharmacists, and care coordinators while preserving clinician autonomy. Our implementation strategy closely aligns with the recently published CKM Guideline, which identifies interdisciplinary care, coordinated implementation of guideline-directed therapies, comprehensive medication management, and mitigation of healthcare fragmentation as central principles of CKM management.^49^ By integrating explainable implementation prioritization with clinician-readable recommendations and multidisciplinary workflows, CHAPERONE-CKM operationalizes many of these guideline recommendations within a Learning Health System framework and provides a potential mechanism for translating guideline-directed care into routine clinical practice

### Explainability, Learning Health Systems, and Responsible AI

Learning Health Systems require continuous interaction between clinical care, data analytics, and quality improvement. CHAPERONE-CKM was designed to support longitudinal electronic health record surveillance, multidisciplinary review, and iterative reassessment as patient status evolves. Transparent explanations identifying the principal digital biomarkers underlying each recommendation enhance clinician confidence, facilitate multidisciplinary communication, and maintain physician oversight.

Successful implementation requires transparency, governance, reproducibility, and continuous monitoring in addition to predictive performance. Consistent framework behavior across synthetic demographic and clinical subgroups provides preliminary support for continued evaluation; however, synthetic validation cannot capture healthcare access, social determinants of health, clinician practice variation, or medication affordability. Consequently, external validation should examine calibration, implementation fidelity, workflow integration, clinician adoption, and equity across diverse healthcare settings.

### Relationship to Existing Risk Models

Traditional cardiovascular risk equations, including PREVENT, estimate future cardiovascular risk but were not developed to coordinate multidisciplinary CKM care. CHAPERONE-CKM complements rather than replaces these tools by integrating multidomain digital biomarkers with explainable implementation prioritization and clinician-readable recommendations that support population health management and value-based care.

### Strengths and Limitations

Strengths include evaluation within a large synthetic cohort encompassing the CKM continuum, incorporation of multidomain demographic, laboratory, imaging, therapeutic, and healthcare utilization variables, and deliberate separation of synthetic data generation from framework evaluation to minimize deterministic relationships. The emphasis on implementation science, explainability, Learning Health Systems, and responsible AI extends beyond conventional assessments of predictive performance.

Several limitations warrant consideration. The framework was evaluated exclusively in a synthetic environment and therefore cannot establish clinical effectiveness, real-world predictive performance, or healthcare equity. Workflow simulations demonstrate implementation feasibility but not improvements in patient outcomes, clinician adoption, healthcare utilization, or cost-effectiveness. In addition, synthetic populations cannot fully reproduce biological complexity, social determinants of health, healthcare access, or regional practice variation. External validation and prospective implementation studies remain essential before clinical deployment.

### Conclusions

CHAPERONE-CKM is an explainable, clinician-supervised implementation framework designed to integrate multidomain digital biomarkers into coordinated CKM population health management. Synthetic validation demonstrated coherent implementation behavior, transparent clinical reasoning, stable subgroup performance, and compatibility with Learning Health System principles. These findings represent an early translational milestone supporting progression to external validation, silent electronic health record deployment, prospective registries, and pragmatic implementation trials. More broadly, this work supports a shift in healthcare AI from isolated prediction toward explainable implementation frameworks that strengthen multidisciplinary care, facilitate guideline-directed therapy, and advance equitable, value-based CKM management.

## Future Directions

Future work will focus on external validation using geographically diverse electronic health record datasets to evaluate calibration, implementation fidelity, clinician adoption, workflow integration, and performance under real-world conditions while monitoring for calibration drift and unintended bias. Subsequent prospective implementation studies should determine whether explainable clinician-supervised AI improves multidisciplinary referral, optimization of guideline-directed medical therapy, patient-reported outcomes, healthcare utilization, and cost-effectiveness. Particular emphasis should be placed on health equity by evaluating implementation across rural communities, socioeconomically disadvantaged populations, and historically underserved racial and ethnic groups to ensure that AI-supported CKM care promotes equitable access to evidence-based therapies and value-based healthcare delivery.

## Take-Home Message

The principal contribution of this investigation is not the demonstration of a high-performing prediction model, but the development of an explainable implementation framework designed to support coordinated CKM care. Synthetic data validation establishes a foundation for external validation and prospective implementation research while emphasizing transparency, clinician oversight, multidisciplinary collaboration, and continuous learning as essential components of responsible artificial intelligence in cardiovascular population health.

## Clinical Perspective

### What Is New?

CHAPERONE-CKM is presented as an explainable, clinician-supervised implementation framework designed to support multidisciplinary cardiovascular–kidney– metabolic population health management rather than as a stand-alone predictive algorithm. Synthetic data validation demonstrated internally coherent framework behavior, transparent clinician-readable explanations, implementation readiness, and compatibility with Learning Health System principles.

### What Are the Clinical Implications?

Explainable clinician-supervised artificial intelligence may help identify patients who would benefit from coordinated multidisciplinary CKM care, facilitate implementation of guideline-directed medical therapy, and support population health management while preserving clinician oversight. Prospective studies are required to determine whether this implementation strategy improves clinical outcomes, health equity, healthcare utilization, and value-based performance in routine practice.

### Ethical Approval

The broader CHAPERONE-CKM study was conducted in accordance with the Declaration of Helsinki, International Council for Harmonisation Good Clinical Practice guidelines, and applicable institutional and regulatory requirements. The protocol was reviewed and approved by Elemental Independent Review Board, 401 Ryland St., STE 200-A, Reno, NV 89502, USA; OHRO Submission Number 370344. Written informed consent was obtained from participants before completion of Phase Ia procedures. For Phase Ib, all records were synthetic and contained no identifiable human data; therefore, separate informed consent was not required. ClinicalTrials.gov identifier: NCT07403669.

**FIGURE 1:**
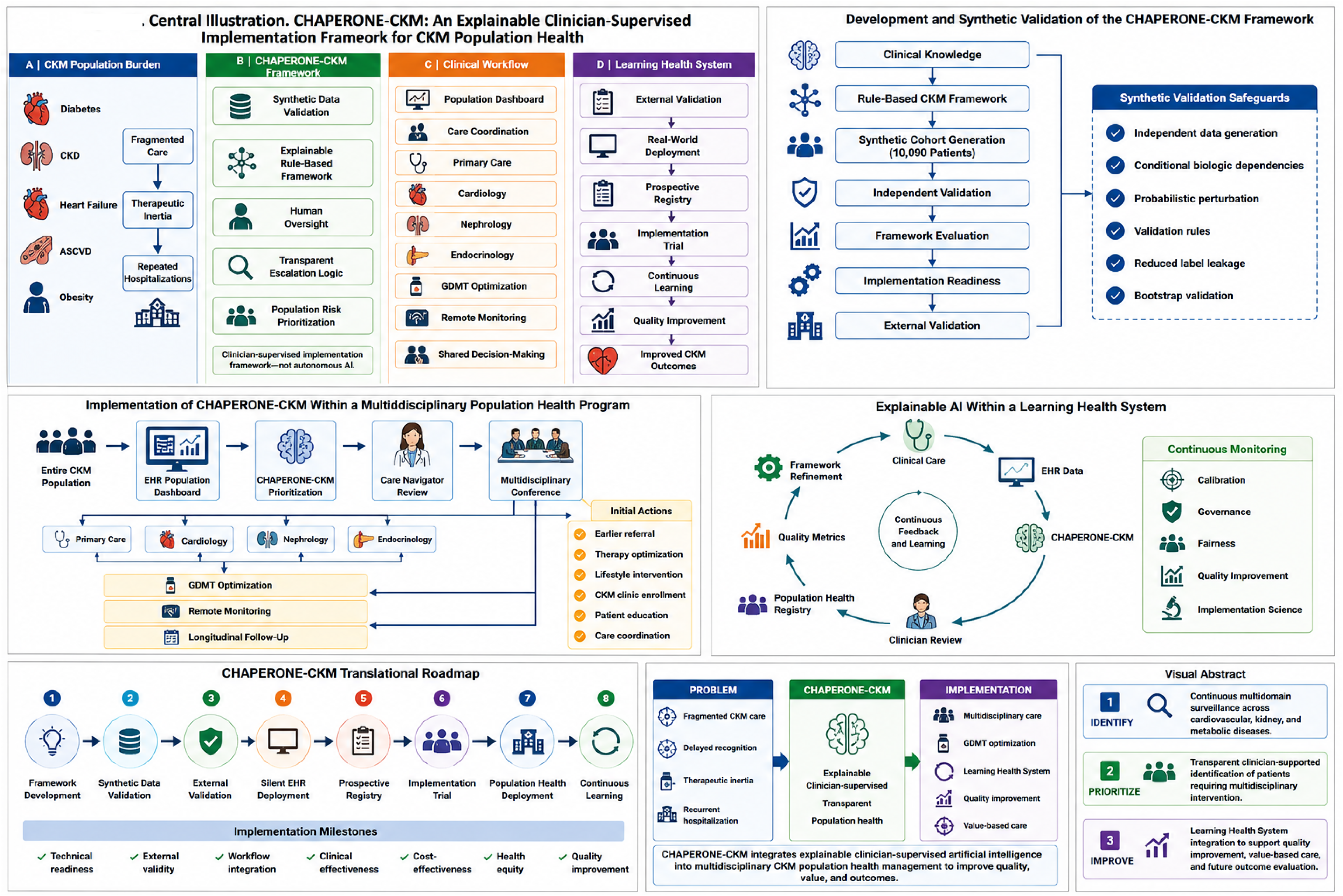
CENTRAL ILLUSTRATION: Explainable AI, CKM Framework, and Implementation Model of CHAPERONE. Uploaded

**Figure 2:**
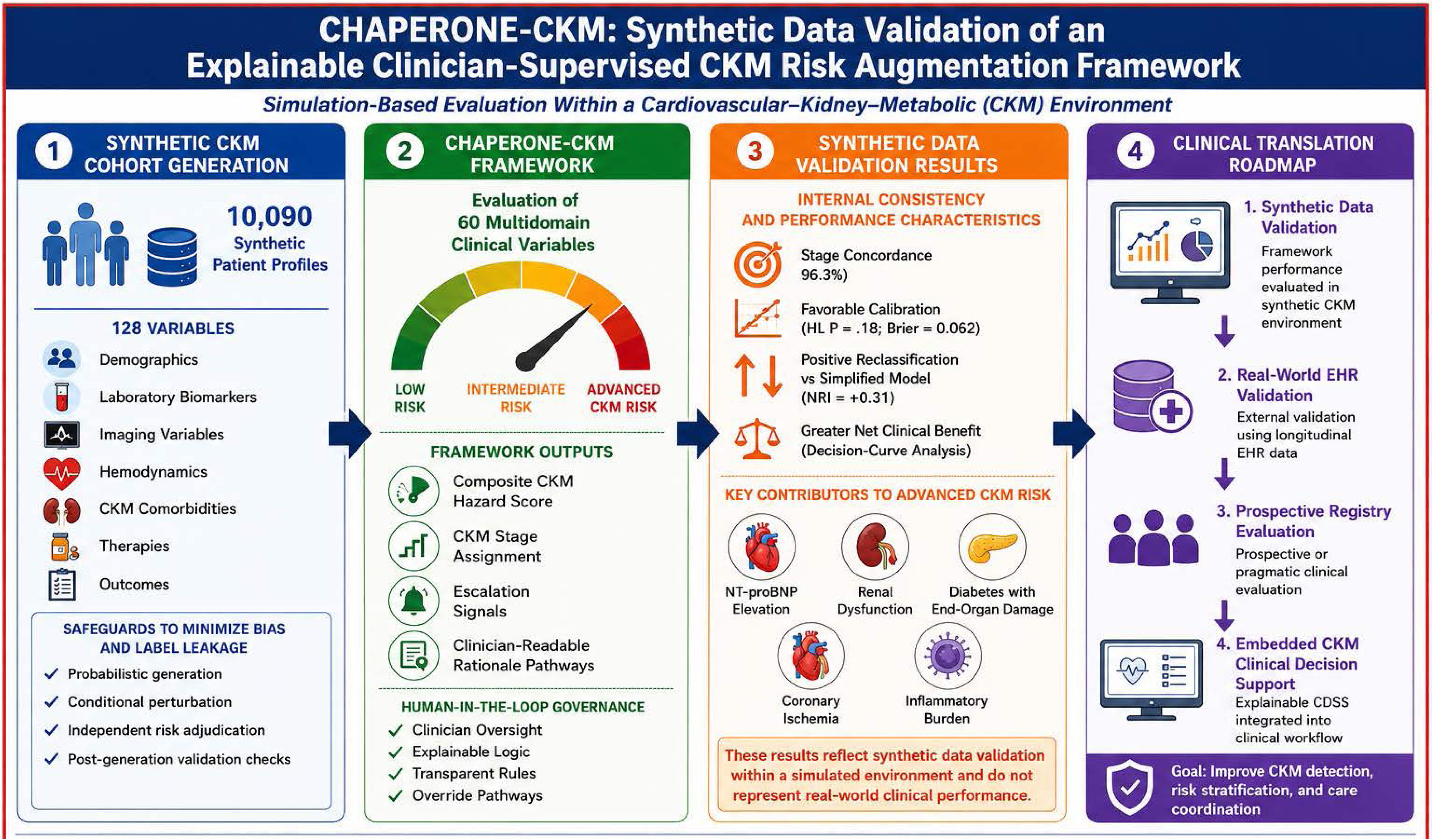
Synthetic Data Validation, Supervised Risk Augmentation Framework and Clinical Translation Roadmap. Uploaded

## Data Availability

The complete software framework and implementation engine constitute proprietary intellectual property. Synthetic data specifications, variable definitions, and analytical workflows sufficient to reproduce the published analyses will be made available upon reasonable request while protecting proprietary algorithms.

## Data Sharing Statement

The synthetic dataset generated for this study was created exclusively for methodological development and validation of the CHAPERONE-CKM implementation framework. No identifiable patient information or protected health information was used in the analyses reported in this manuscript. The computational framework, synthetic data generation methodology, variable definitions, statistical analysis plan, and implementation logic are described, to the extent possible in the manuscript and Supplementary Appendix while protecting intellectual Property.

## Funding

The CHAPERONE CKM study number AVDH 008; CHAPERONE -CKM trial is sponsored and funded by Aventyn, Inc, and was also made possible by a partial grant from Arizona Commerce Authority Partnership for Economic Innovation

The CHAPERONE CKM platform includes non-proprietary and open-source components used under appropriate licenses

## Conflict of Interest

Krishnaswami Vijayaraghavan is involved in the scientific development of the CHAPERONE CKM platform. This role has been disclosed and managed in accordance with institutional policies. All AI-generated outputs evaluated in this study underwent independent clinician review and adjudication. Navin Govind is the CEO of Aventyn,Inc. The remaining authors declare no competing interests.

## Acknowledgements

Domain experts, Data and technical ream and human evaluation team that included Aditya Vijay, Navin Govind, Arjun Moorthy, Sun Jones, East Valley Endocrinology, Samer Ibrahim, Keyvan Koochek

## Author contributions

Conceptualization: Krishnaswami Vijayaraghavan, Navin Govind, Zaki Lababidi

Methodology: Krishnaswami Vijayaraghavan, Navin Govind, Samer Ibrahim, Keyvan Koochek, Sun Jones

Software: Navin Govind, Aditya Vijay, Arjun Moorthy

Validation: Krishnaswami Vijayaraghavan, Sun Jones, Samer Ibrahim, Keyvan Koochek, Devendra Wadwekar

Formal Analysis: Krishnaswami Vijayaraghavan, Aditya Vijay, Arjun Moorthy, Navin Govind

Investigation: Krishnaswami Vijayaraghavan, Aditya Vijay, Arjun Moorthy, Lakshmi Nair, Sun Jones, Zaki Lababidi

Resources: Navin Govind, Randal Schulhauser, Edgar Lerma, Marcus Stahlberg, Dinesh Kalra, Patrick Dunn, Kevin Shah, Jordi Livi, Dinesh Kalra, Krishnaswami Vijayaraghavan

Data Curation: Aditya Vijay, Sun Jones, Devendra Wadwekar, Samer Ibrahim

Writing – Original Draft: Krishnaswami Vijayaraghavan, Arjun Moorthy, Aditya Vijay, Navin Govind

Writing – Review & Editing: All authors

Visualization: Krishnaswami Vijayaraghavan, Aditya Vijay, Arjun Moorthy, Navin Govind Supervision: Krishnaswami Vijayaraghavan, Navin Govind

Project Administration: Krishnaswami Vijayaraghavan, Aditya Vijay, Sun Jones, Devendra Wadwekar

## Funding Acquisition

Navin Govind

All authors reviewed and approved the final manuscript and agree to be accountable for all aspects of the work.

## Non-Standard Abbreviations and Acronyms

CKM: Cardiovascular–Kidney–Metabolic
ASCVD: Atherosclerotic Cardiovascular Disease
GDMT: Guideline-Directed Medical Therapy
LHS: Learning Health System
CFIR: Consolidated Framework for Implementation Research
RE-AIM: Reach, Effectiveness, Adoption, Implementation, Maintenance
CDS: Clinical Decision Support
EHR: Electronic Health Record
AI: Artificial Intelligence
NT-proBNP: N-terminal Pro-B-type Natriuretic Peptide

## Notes

### Competing Interest Statement

The authors have declared no competing interest.

### Clinical Trial

NCT07403669

